# Heterologous ChAdOx1 nCoV-19 and BNT162b2 prime-boost vaccination elicits potent neutralizing antibody responses and T cell reactivity

**DOI:** 10.1101/2021.05.30.21257971

**Authors:** Rüdiger Groß, Michelle Zanoni, Alina Seidel, Carina Conzelmann, Andrea Gilg, Daniela Krnavek, Sümeyye Erdemci-Evin, Benjamin Mayer, Markus Hoffmann, Stefan Pöhlmann, Alexandra Beil, Joris Kroschel, Bernd Jahrsdörfer, Hubert Schrezenmeier, Frank Kirchhoff, Jan Münch, Janis A. Müller

## Abstract

Heterologous COVID-19 vaccination regimens combining vector- and mRNA-based vaccines are already administered, but data on solicited adverse reactions, immunological responses and elicited protection are limited. We aimed to evaluate the reactogenicity, humoral and cellular immune responses towards different SARS-CoV-2 variants after a heterologous ChAdOx1 nCoV-19 BNT162b2 prime-boost vaccination and analyzed a cohort of 26 individuals aged 25-46 (median 30.5) years that received a ChAdOx1 nCoV-19 prime followed by a BNT162b2 boost after an 8- week interval. Self-reported solicited symptoms after ChAdOx1 nCoV-19 prime were in line with previous reports and less severe after the BNT162b2 boost. Antibody titers increased significantly over time resulting in strong neutralization titers two weeks after the BNT162b2 boost. Neutralizing activity against the prevalent strain B.1.1.7 (Alpha) and immune-evading VOC B.1.351 (Beta) was ∼4-fold higher than in individuals receiving homologous BNT162b2 vaccination. No difference was seen in neutralization of VOI B.1.617 (Kappa). In addition, the heterologous vaccination induced CD4+ and CD8+ T cells reactive to SARS-CoV-2 spike peptides of all analyzed variants; Wuhan-Hu-1, B.1.1.7, B.1.351, and P.1 (Gamma). In conclusion, heterologous ChAdOx1 nCoV-19 / BNT162b2 prime-boost vaccination regimen is not associated with serious adverse events and results in a potent humoral immune response and elicits T cell reactivity. Variants B.1.1.7, B.1.351 and B.1.617.1 are potently neutralized by sera of all participants and reactive T cells recognize spike peptides of all tested variants. These results suggest that this heterologous vaccination regimen is at least as immunogenic and protective as homologous vaccinations.

## Introduction

The first cases of the coronavirus disease 2019 (COVID-19) were reported to the World Health Organization on December 31^st^ 2019 (1), and within 93 days the causative severe acute respiratory syndrome coronavirus 2 (SARS-CoV-2) had infected over 1 million people worldwide (2). Only 250 days later, the first person received a COVID-19 vaccine outside a clinical trial, and vaccinations are now considered a key strategy for ending the pandemic(3). Approved vaccines include the adenovirus-based ChAdOx1 nCoV-19 (Vaxzevria, AstraZeneca) and mRNA-based BNT162b2 (Comirnaty, BioNTech/Pfizer), which induce humoral and cellular immunological responses (4–8), showed high efficacy in clinical trials (9, 10) and a high degree of protection from COVID-19 in real-world settings(11, 12). However, the occurrence of rare thrombotic events with thrombocytopenia after ChAdOx1 nCoV-19 vaccinations, especially in individuals younger than 60 years, associated with the generation of auto-platelet factor 4 antibodies, halted vaccination of this group with ChAdOx1 nCoV-19 in some countries (13–15). As a consequence, several public health agencies now recommend that boost vaccination of individuals already primed with ChAdOx1 nCoV-19 is carried out in a heterologous regimen with an mRNA vaccine (16). In others, it is not applied due to limited data on safety and immunologic responses elicited by such a regimen. Two recent studies and preprints indicate that such a heterologous schedule is associated with more severe (17) or similar (18–20) solicited symptoms, respectively. Similarly, one published article and upcoming preprints suggest that robust immune responses are elicited (18–22), but there is limited knowledge about T cell responses or protection against the variants of concern (VOC) by heterologous vaccination regimens (20, 22). Here, we studied a cohort of 26 individuals (16 female, 10 male; median age 30.5, range 25-46) (Table 1) who received ChAdOx1 nCoV-19 prime and, due to changing recommendations in Germany, (16) a BNT162b2 boost vaccination with a 56 day interval and evaluated solicited adverse reactions, humoral and cellular immune responses against several spike variants.

**Table 1.**
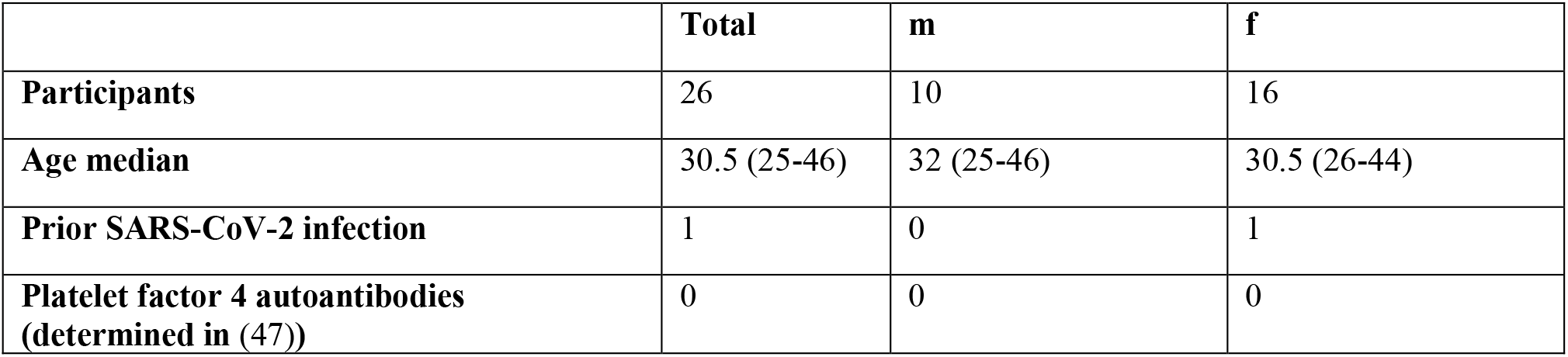
Study participants:

## Results

Reactogenicity following prime and boost vaccination was evaluated by all study participants by self-reporting of solicited local and systemic symptoms according to a standardized questionnaire. Symptom severity (mild, moderate, severe) and duration (<1 h, few h, ∼1 day, > 1 day) is reported for each individual participant (Figure S1A) and percentage of participants (Figure 1A,B).

**Fig. 1.**
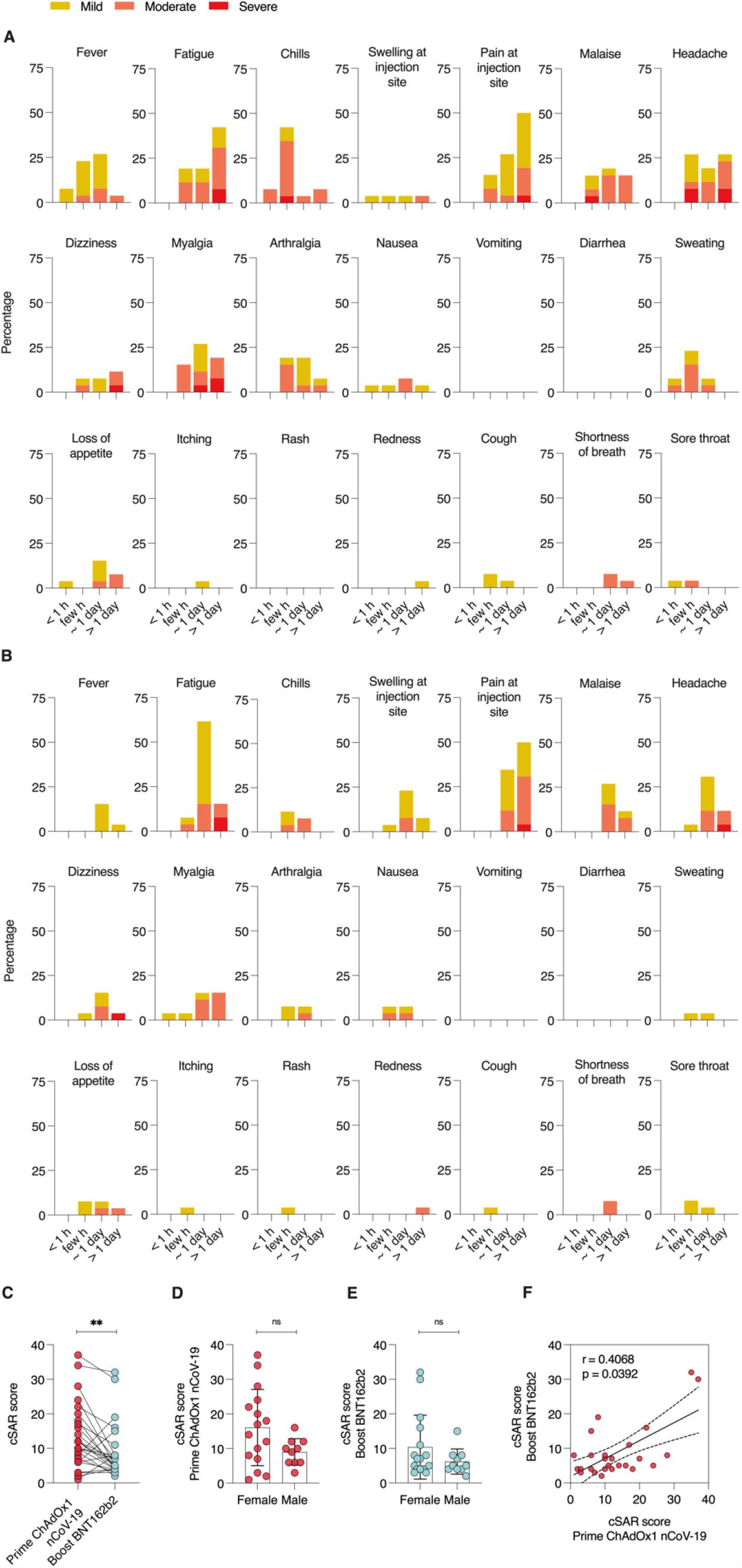
Solicited adverse reactions following ChAdOx1 nCoV-19 prime and BNT162b2 boost vaccination. Percentages of participants with individual symptoms following prime (A) or boost (B) vaccination. Severity is graded on a scale of 1-2 (for some symptoms) or 1-3 (for most), according to Common Terminology Criteria for Adverse Events (US Department of Health and Human Services, Version 4.03) (23). (C) Cumulative solicited adverse reaction (cSAR) scores of all participants following prime and boost vaccination. For calculation of cSAR scores, symptom gradings are summed and an additional score point is added for symptoms lasting more than 24 h. Analysis of cSAR scores by (D, E) participant gender, and (F) comparison between cSAR scores following prime and boost vaccination. The SARS-CoV-2 convalescent individual was excluded in all statistical analyses. Paired t-test; ns not significant; ** p < 0.01

Both, prime and boost vaccination, induced mild to moderate solicited adverse reactions in most participants with 88.4% (23/26) reporting at least one mild or moderate symptom following prime; 23/26 (88.4%) and 21/26 (80.8%) reporting at least one mild or moderate symptom following boost vaccination (Figure 1A,B). Most common symptoms after prime vaccination with ChAdOx1 nCoV-19 were pain at the injection site (92.3%), fatigue (80.8%), headache (73.1%), chills (61.5%), myalgia (61.5%) and fever (61.5%). Following boost vaccination with BNT162b2, most participants again reported pain at the injection site (84.6%) and fatigue (84.6%), but chills (19.2%), myalgia (38.5) and fever (19.2%) were less common. 23% of participants (6/26) reported at least one severe symptom following prime, 15.4% (4/26) after boost. Fatigue (7.7%) and headache (15.4% for prime, 3.8% for boost) were amongst symptoms reported as severe for both doses, while myalgia was reported as severe by 11.5% of participants following prime, but none after boost.

Comparing cumulative solicited adverse reaction (cSAR) scores, reactogenicity following prime with ChAdOx1 nCoV-19 was significantly (p = 0.008) higher than following boost with BNT162b2 (cSAR score median 11 and 6 respectively, Figure 1C). Individually, most participants (19/26, 73.07%) had milder reaction to boost compared to prime. 6/26 (23.07%) of participants described more severe reactions to boost vaccination (Figure S1B). A trend towards higher cSAR scores reported by female participants was seen for both boost and prime vaccinations (Figure 1D,E). No correlation was observed between reactogenicity and age (Figure S1C,D). Individual reactogenicity towards prime and boost vaccination showed a weak but significant correlation (Figure 1F, p = 0.039).

We collected sera from participants 2 days (−2) or on the same day (0) before vaccination, and at days 15 – 16, 30 – 37, and 53 – 57 after ChAdOx1 nCoV-19 prime, and days 6 - 11 and 14 – 19 after BNT162b2 boost (64 – 65 or 72 – 73 after prime, respectively) to determine antibody responses (Figure 2). Already 15-16 days after prime, 19/25 (76%) participants showed detectable anti-SARS-CoV-2-spike-IgG levels and 17/25 (65%) detectable IgA levels (Figure 2A,B). IgG levels peaked after 30 - 37 days and were detectable in 24/25 (96%) participants. Until days 53 – 57, IgG levels slightly decreased, consistent with previous results after single ChAdOx1 nCoV-19 dose (5, 6). IgA values were highest already at days 15-16 and became undetectable in 24 (92%) participants at days 53 - 57. Notably, only 6 - 11 days after the BNT162b2 boost, IgG was detectable in all (100%) and IgA in 23 (92%) of 25 participants. Until day 14-19 after boost (72- 73 post ChAdOx1 nCoV-19), IgG and IgA were detectable in all participants. This corresponds to an at least 3.7-fold increase in median IgG levels from pre-boost to 2 weeks post-boost. We next quantified cumulative anti-SARS-CoV-2-spike-IgM and IgG concentrations and detected median antibody levels of 3.39 (range 0-2,126) units per ml (U/ml) 15-16 days after prime vaccination in 22/25 (88%) participants (Figure 2C). From days 30 – 37 on, IgM and IgG were detected in all participants and medians continuously increased to 28 (1.86-1,436) and 63.9 (4.27-1,005) U/ml after days 30 - 37 or 53 - 57, respectively. After BNT162b2 boost, titers increased 134-fold to 8,614 (126 – 24,831) at days 6 – 11 and 135-fold to 8,815 (1,206 – 19,046) 14 - 19 days after the second dose. Strikingly, the resulting titers were 8.1-fold higher than those determined for sera obtained after 13-15 days of a homologous BNT162b2 boost (individuals with median age 41 (25- 55); median titers 1,086; range 498-3,660; Appendix Table 1). Cumulative IgM/G titers correlated with IgG titers at each timepoint analyzed post prime (Figure S2, Appendix Table 2).

**Fig. 2.**
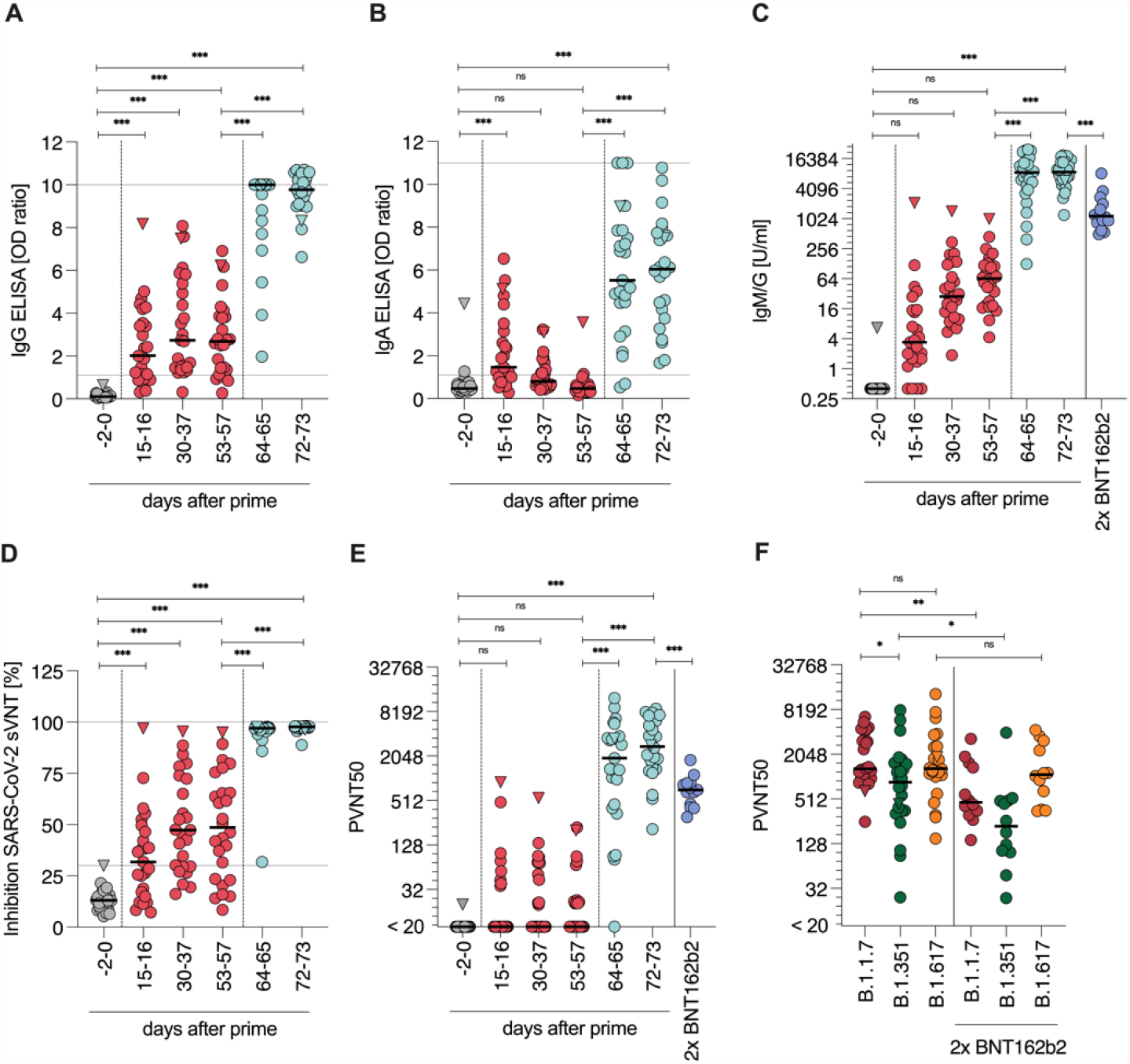
Humoral response. Quantification of anti-SARS-CoV-2 S1 spike domain (A) IgG and (B) IgA titers. (C) Quantification of anti-SARS-CoV-2 spike IgG and IgM responses as units per ml (U/ml) by immunoassay. (D) SARS-CoV-2 surrogate virus ACE2 neutralization test. (E) VSV-based B.1.1.7 SARS-CoV-2 spike pseudovirus neutralization assay. (F) VSV-based B.1.1.7, B.1.351 and B.1.617.1-SARS-CoV-2 spike pseudovirus neutralization assay. Titers expressed as serum dilution resulting in 50% pseudovirus neutralization (PVNT50). Triangle indicates SARS-CoV-2 convalescent individual, who was excluded from all statistical analyses. Grey symbols indicate datapoints pre-vaccination, red datapoints indicate datapoints after prime and light-blue after boost vaccination. Dark-blue indicates samples of participants with homologous BNT162b2 prime-boost regimen. Dashed horizontal lines indicate upper and lower limit of detection/cut-off, respectively. Dashed vertical lines indicate prime and boost vaccination. Longitudinal antibody measurements were analyzed by means of a mixed linear regression model. Mann-Whitney-U test compares ChAdOx1 nCoV-19 and BNT162b2 titers *** p < 0.0001, ** p < 0.001, * p < 0.05, ns not significant

Sera were also evaluated for their potential to inhibit SARS-CoV-2-spike-receptor binding domain/ACE2 interaction using a surrogate virus neutralization test (sVNT) (Figure 2D). 15-16 days after ChAdOx1 nCoV-19 administration 13/25 (52%) participant sera showed ACE2 neutralizing activity, correlating significantly with IgG and IgM/G titers (Figure S2, Appendix Table 2). Median neutralization activity of the positive sera was 46% (range 32-97%). Until days 53-57, the number of participants with neutralizing sera increased to 19/26 (73%) and the median ACE2 neutralization to 62% (range 32-95%), again in correlation with IgG and IgM/G values (Figure S2, Appendix Table 2). After BNT162b2 boost, all participants showed potent neutralization with a median of 97% (range 32-98%) after 6-11, and 98% (range 89-98%) after 14- 16 days suggesting a strong and functional antibody response after heterologous vaccination in all participants.

The potency of neutralizing activity was further quantified using vesicular stomatitis virus (VSV)- based pseudoviruses carrying the SARS-CoV-2 spike protein of the most prevalent SARS-CoV-2 B.1.1.7 (Alpha) variant. This system faithfully recapitulates SARS-CoV-2 entry into cells and its inhibition(24–26). 15-16 days after ChAdOx1 nCoV-19 prime, neutralizing titers ranging from 36-906 were detectable in 8/25 (32%) participants (Figure 2E). The number of participants with detectable neutralization increased to maximum in 12/25 (48%) individuals at days 30-37 with a median neutralization titer of 74 (range 20-552) in responders, which slightly decreased until days 53-58. Two weeks after the BNT162b2 boost, neutralizing titers were detected in all participants with a median titer of 2,744 (range 209-8,985). Of note, while for some individuals the titers further increased from week 1 to week 2 after BNT162b2 boost, other individuals plateaued at titers > 1,000 (Figure S3). At all time points, neutralizing activity correlated with IgG or IgM/G titers (Figure S2, Appendix Table 2). Remarkably, the median titer of these individuals was 3.9-fold higher than the median titer of 14 individuals vaccinated with BNT162b2 in a homologous regimen (709; range 305-1,806) suggesting a stronger humoral protection after a heterologous vaccination. Of note, a SARS-CoV-2 convalescent individual (triangle symbol) showed a strong response after the first dose in all assays, high IgG, IgA or IgM/G values, most effective ACE2- neutralization and a high neutralization titer of 906 15 – 16 days after prime that decreased over the days to 201 at day 53-57 (Figure 2A-E).

Additionally, we evaluated the neutralizing activities of sera obtained 2 weeks post full vaccination against the immune evading variants B.1.351 (Beta) and B.1.617.1 (Kappa). Pseudovirus entry driven by B.1.351 spike was neutralized with 2-fold lower potency (p < 0.05) compared to B.1.1.7 spike. However, it was still entirely blocked at higher doses with a median titer of 1,297 (range 252 - 6,523). Neutralization of the B.1.617.1 spike was not reduced compared to B.1.1.7 spike (median titer of 1,309; range 150 – 13,252) (Figure 2F). Sera of individuals vaccinated with homologous BNT162b2 showed lower neutralizing titers against all spike variants tested (Figure 2F), suggesting stronger humoral protection after a heterologous vaccination also against VOCs. Notably, no correlation of neutralizing antibody titre (PVNT50) with sex or age was observed for either cohort (Figure S4).

To evaluate cellular immunity, we isolated peripheral blood mononuclear cells from blood samples provided by 21/26 participants before ChAdOx1 nCoV-19 prime, and 6–11 days post BNT162b2 boost and 14-19 days post boost (72-73 days post prime), considered as full vaccination according to the vaccination schedule. Cells were exposed to SARS-CoV-2 spike-spanning peptide-pools and analyzed for intracellular cytokines TNFα, IFNγ, and IL-2 to determine spike-specific CD4^+^ and CD8^+^ T cell responses (Figure 3, S5, S6). 6-11 days after boost, 45-65% of participants already showed IFNy, TNFα, or IL-2 producing CD4^+^ T cells upon stimulation with Wuhan-Hu-1 SARS-CoV-2 spike peptides (Figure 3A). After full vaccination, CD4^+^ T cells producing IFNγ (median 0.055, range 0.018-0.168), IL-2 (median 0.055, range 0-0.134) or TNFα (median 0.057; range 0.01 – 0.193) were significantly increased upon stimulation as compared to the pre-vaccination baseline. Spike reactive CD4^+^ T cells were detected in 63-95% suggesting that most individuals developed a robust spike-specific T helper 1 (TH1) CD4^+^ T cell response post BNT162b2 boost. In addition, we observed an increase of spike-specific CD8^+^ T cells predominantly producing IFNγ (median 0.092, range 0-0.665) and TNFα (median 0.055, range 0 – 0.375) compared to pre-vaccination baseline samples in all previously uninfected participants (Figure 3A). Levels of spike- specific CD8^+^ T cells producing IL-2 (median 0.01, range 0-0.052) were lower, which is in agreement with responses after homologous BNT162b2 vaccination (7). We then compared the response to SARS-CoV-2 spike variants B.1.1.7, B.1.351, and P.1 (Gamma) at day 6-11 after boost, and found that both CD4^+^ and CD8^+^ T cells showed IFNγ responses to all spike variants (Figure 3B), however less pronounced for TNFα and IL-2 (Figure S6 C,D). CD4^+^ T cell reactivity towards B.1.351 spike was slightly decreased and CD8^+^ T cell reactivity towards B.1.1.7 spike was slightly elevated, but overall a robust cellular immune response against all three spike variants was found in most participants. Of note, reactivity of the CD4^+^ and CD8^+^ T cells of the convalescent individual did not increase after vaccination (Figure 3A), indicating pre-existing cellular immunity. Overall, these findings show a robust humoral and cellular immune response after heterologous vaccination.

**Fig. 3.**
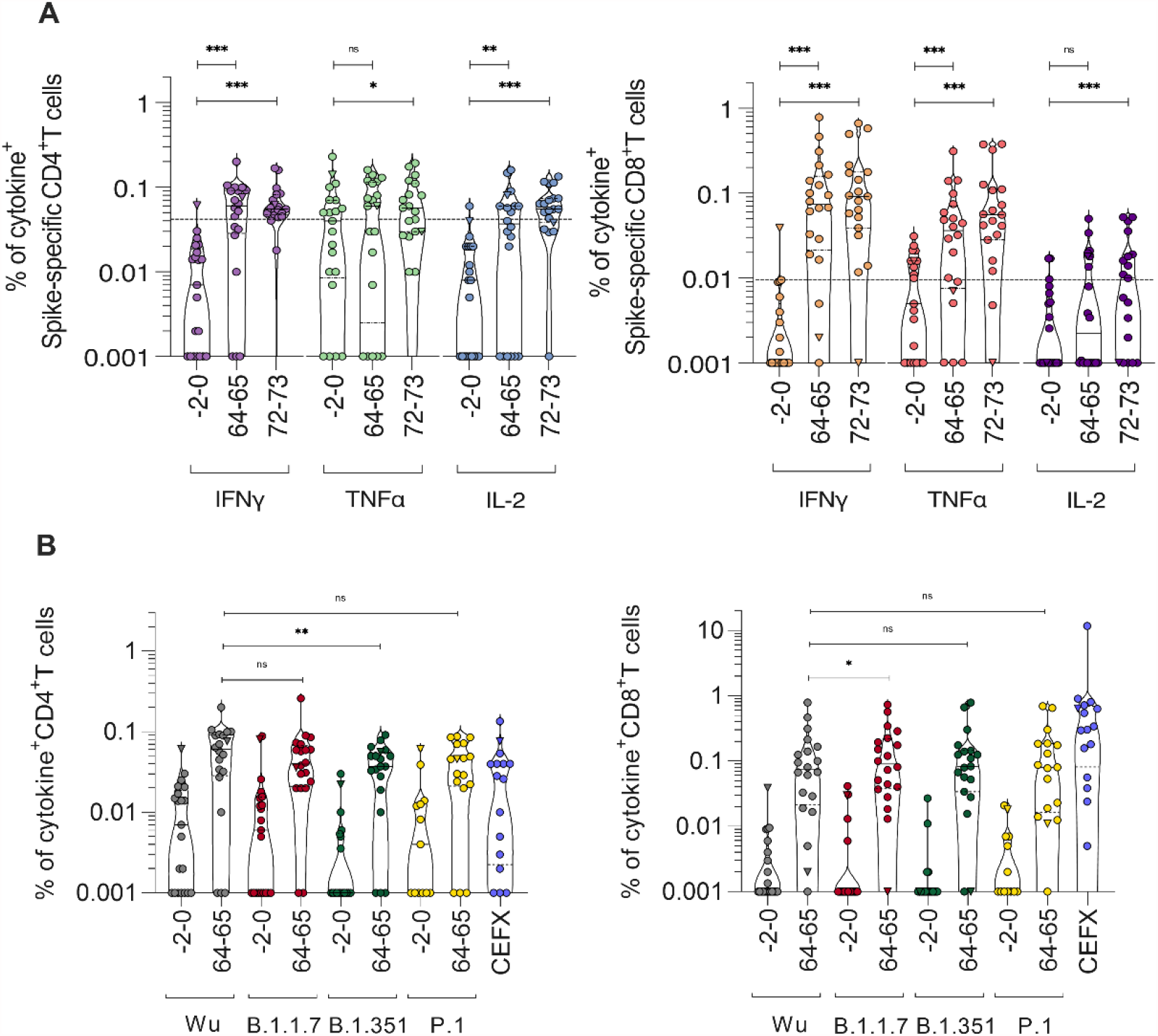
SARS-CoV-2 spike-specific CD4^+^ and CD8^+^ T cell responses. PBMCs from day -0-2 pre- and day 64-65 or 72-73 post prime vaccination (1-2 weeks post boost) of study participants were stimulated with (A) SARS-CoV-2 Wuhan-Hu-1 spike peptide-pool and cytokine production determined by flow cytometry. (B) PBMCs from day -0-2 pre and day 64-65 post prime were stimulated with SARS-CoV-2 spike peptide pools derived from B.1.1.7, B.1.351, or P.1. or of epitopes of different infectious agents (CEFX) and compared with Wuhan-Hu-1 from (A). Cytokine^+^ T cells were background-corrected for unstimulated cells. Values lower than median plus one standard deviation of pre-vaccination (0.04% for CD4, 0.01% for CD8) were considered negative. Triangle symbol indicates SARS-CoV-2 convalescent individual, where cytokine release was already high in absence of stimulation. Wilcoxon matched-pairs signed rank test compares cytokine-positive cells before and after vaccination (1-2 weeks post-boost) or upon stimulation with different SARS-CoV-2 spike variants, *** p < 0.0001, * p < 0.05.

## Discussion

Based on the regulatory approvals for ChAdOx1 nCoV-19 and mRNA vaccines, the interval between prime and boost vaccinations ranges between 4 -12 weeks (27–29). For ChAdOx1 nCoV- 19, a 12 week interval has been shown to result in stronger immune responses (30), most likely because the immunity against the vector wanes. Accordingly, e.g. in Germany heterologous vaccinations are currently typically performed after 12 weeks. Existing vector immunity, however, is irrelevant in the context of a mRNA boost vaccination, on which basis our cohort received the boost after 8 weeks. This heterologous ChAdOx1 nCoV-19/BNT162b2 vaccination elicited strong IgM/G and IgA responses, neutralizing activities and T cell responses in all previously uninfected participants, while solicited adverse reactions to vaccination were as expected for a prime ChAdOx1 nCoV-19 vaccination and reduced following heterologous BNT162b2 boost.

A previous study showed that a heterologous vaccination schedule with 4-week interval results in stronger reactogenicity after boost (17), whereas a published and a pre-printed studies with an 8- to 12-week interval did not confirm this effect (18–20). We did not directly compare different vaccination schemes. Thus, we cannot draw definitive conclusions on differences, which might also depend on cohort age (6). With an 8-week interval, we observed an overall milder reactogenicity following heterologous boost with BNT162b2 than after initial prime vaccination with ChAdOx1 nCoV-19 and no serious adverse events, arguing for the safety of this regimen in young adults.

Our immunological data suggest that a heterologous vector-based/mRNA prime-boost schedule is highly effective in preventing COVID-19, as neutralizing antibody levels correlate with immune protection from symptomatic SARS-CoV-2 infection (31) and CD8^+^ T cell responses have been associated with a mild disease course (32, 33). Endpoint neutralizing antibody titers determined 2 weeks post boost were slightly higher than those detected upon homologous BNT162b2 vaccinations (Figure 2 C,E). This is in agreement with results from pre-printed studies (20–22) and suggests that ChAdOx1 nCoV-19 / BNT162b2 prime-boost vaccination elicits potent immune responses (18, 19). However, results might also be influenced by the young cohort age. Factors contributing to this high degree of immunogenicity might be the circumvention of vector immunity. The BNT162b2 encoded spike sequence contains a two-proline mutation not present in ChAdOx1 nCoV-19, which fix spike in a pre-fusion confirmation (10, 34). It is tempting to speculate that altered spike conformations between the vaccinations may be beneficial for effective neutralizing responses.

Neutralizing activity towards VOC B.1.351, previously reported to show partial evasion of vaccination-induced antibodies (24, 35, 36), was slightly decreased compared to B.1.1.7 following heterologous vaccination, again in agreement with recently pre-printed data (22). However, the titer was still higher than for B.1.1.7 after two doses of BNT162b2. Neutralization of emerging B.1.617.1, which appears to be the most efficiently neutralization-evading strain of the B.1.617 lineage(37), was not reduced compared to B.1.1.7. Whether these immunological findings translate into effective general protection from VOCs in real-life setting remains to be determined, however neutralizing antibody titers have been described to be a good predictor of protective vaccine efficacy (31) and the substantial neutralization capacity against the most prevalent and evading virus variants is encouraging.

Secretory IgA responses at the mucosal site of SARS-CoV-2 entry are of particular interest with regard to prevention of virus transmission and (re-)infection (38). We detected a general increase in serum IgA levels with strong variation between participants, suggesting mucosal protection shortly after vaccination. However, IgA levels decreased over time after prime vaccination, and future studies, especially assessing IgA and secretory IgA levels and persistence at mucosal entry sites after boost are warranted.

In almost all participants SARS-CoV-2 specific CD8^+^ or CD4^+^ T cells were detected 2 weeks after full vaccination. These effects were similar to those reported after a single ChAdOx1 nCoV-19 dose and after homologous BNT162b2 vaccination (5, 7). This suggests that T cell responses are similarly effective after heterologous vaccination. Importantly, T cell reactivity was potent against most prevalent VOC B.1.1.7 and most humoral immunity-evading VOCs B.1.351 and P.1. This is in line with previous findings that variants escaping humoral immunity are still recognized by reactive T cells indicating broad protection from SARS-CoV-2 on a cellular level (10, 39), also after heterologous vaccination.

In line with previous results, in an individual participant who was previously tested SARS-CoV-2 positive, a single prime vaccination dose already elicited strong antibody responses (40, 41). In this case, the observed neutralizing titers 2 weeks after prime were as high as the median titer of those receiving the homologous BNT162b2 vaccination. However, titers (IgM/G) further increased 8-fold after boost, suggesting that prime-boost might provide more potent and longer lasting protection also in convalescent patients.

In conclusion, heterologous vaccination schedule of ChAdOx1 nCov-19 prime, followed by BNT162b2 boost after 8 weeks for participants with a median age of 30.5 years was, within our cohort, tolerable and effective. This regimen provides flexibility for future vaccination strategies and will be useful for vaccine schedules during shortages. The perspective that heterologous vaccination might be superior to homologous regimens should be considered as a potential strategy to elicit particularly strong immune responses e.g. in immunocompromised, highly exposed individuals or against VOCs in future prime-boost vaccinations and vaccine updates against COVID-19. Similarly, whether other vector- or mRNA-based vaccine combinations or those based on other technologies are as effective needs to be addressed in future studies.

## Materials and Methods

### Study Design

A cohort of 26 individuals aged 25-46 (median 30.5) years that received a ChAdOx1 nCoV-19 prime followed by a BNT162b2 boost after an 8-week interval were analyzed for reactogenicity, antibody responses and T cell reactivity. Participants were enrolled prior to initial vaccination and initial samples taken up to 2 days before prime vaccination. Solicited adverse reactions after both prime and boost vaccination were self-reported and ranked by severity and duration. Blood samples were taken from all participants at several time points throughout the study and analyzed for antibodies and T-cell reactivity against SARS-CoV-2 spike.

### Collection of serum and PBMC samples

Blood samples from individuals were obtained after recruitment of participants and written informed consent as approved by the ethics committee of Ulm university (99/21). Participants received a heterologous vaccination regimen because after their ChAdOx1 nCoV-19 prime vaccination, the German Standing Committee on Vaccination (STIKO) had changed the recommendation for individuals < 60 years of age to receive an mRNA vaccine as boost vaccination to avoid risk of thrombotic complications (13, 16). At days -2/0 before vaccination, days 15-16, 30-37, 53-57 after ChAdOx1 nCoV-19 vaccination, and days 6-11 and 14-19 after heterologous BNT162b2 boost (days 64-65 and 72-73 after ChAdOx1 nCoV-19, respectively), blood was drawn into S-Monovette® Serum Gel (Sarstedt) or S-Monovette® K3 EDTA tubes. Sera from individuals vaccinated twice with BNT162b2 were obtained 13-15 days after the second dose under approval by the ethics committee of Ulm university (31/21); these sera were previously described and re-analyzed for this study(24). Serum Gel collection tubes were centrifuged at 1,500 × g at 20°C for 15 min, aliquoted stored at -20°C until further use. Peripheral blood mononuclear cells (PBMCs) were obtained from EDTA tubes using density gradient centrifugation by Pancoll human (Pan Biotech, Germany), and erythrocytes removed by ACK lysis buffer (Lonza, Walkersville, MD, U.S.A). Mononuclear cells were counted for viability using a Countess II Automated Cell Counter (Thermo Fisher) with trypan blue stain and were cryopreserved in aliquots of up to 1×10^7^ cells in 10% DMSO in heat-inactivated FCS.

### Vaccine reactogenicity

Solicited adverse reactions (SAR) were self-reported by the participants via questionnaire following prime and boost vaccination. Participants were asked to list symptoms, their duration (< 1 h, few hours, one day or more than one day) and severity (mild (grade 1), moderate (grade 2), severe (grade 3). Grading criteria were adapted from the US Department of Health and Human Services CTCEA (Common Terminology Criteria for Adverse Events, v4.03) (23), with grade 1- 2 being considered for some symptoms, grade 1-3 for most. For calculation of cumulative SAR (cSAR) scores, the grades of all symptoms listed were summed up, with an additional score point added for symptoms experienced for more than one day (0-4).

### Determination of antibody titers

IgG and IgA levels in serum were determined by anti-SARS-CoV-2 assay (Euroimmun), an ELISA which detects antibodies against the SARS-CoV-2 S1 spike domain. The assay was performed according to the manufacturer’s instructions. Briefly, serum samples were diluted 10- fold in sample buffer and pipetted into rSARS-CoV-2 spike precoated strips of eight single wells of a 96-well microtiter. After incubation for 60 min at 37°C, wells were washed three times, peroxidase-labeled anti-IgG or anti-IgA added and incubated. After 30 min, three additional washing steps were performed before substrate was added and incubated for 15-30 min in the dark. Thereafter, stop solution was added, and optical density (OD) values measured on a POLARstar Omega plate reader (BMG LABTECH, Ortenberg, Germany) at 450 nm corrected for 620 nm. Finally, OD ratios were calculated based on the sample and calibrator OD values, where a ratio <0·8 was considered to be negative and >1·1 to be positive. To quantify antibody responses, IgG and IgM were measured as units per ml (U/ml) that correlates with the WHO standard unit for the SARS-CoV-2 binding antibody units per ml (BAU/ml). To this end, serum was analyzed using the commercial electrochemiluminescence Elecsys Anti-SARS-CoV-2 S immunoassay (Roche, Mannheim, Germany) by a cobas® e801 immunoassay analyzer according to the manufacturer’s instructions (Roche).

### Surrogate SARS-CoV-2 neutralization test

Prevention of SARS-CoV-2 spike RBD interaction with ACE2 by sera was evaluated by SARS-CoV-2 Surrogate Virus Neutralization Test Kit (GenScript) according to the manufacturer’s instructions. To this end, sera were incubated with a peroxidase-conjugated RBD fragment and the mixture added to a human ACE-2 coated plate, and unbound RBD washed away. Thereafter, substrate was added and the reaction stopped by stopping reagent. ODs at 450 nm were measured at a microplate reader. The inhibition score compared to the negative control was calculated as percentages. Scores <20% were considered negative and scores >20% positive.

### Cell culture

Vero E6 (African green monkey, female, kidney; CRL-1586, ATCC, RRID:CVCL_0574) cells were grown in Dulbecco’s modified Eagle’s medium (DMEM, Gibco) which was supplemented with 2·5% heat-inactivated fetal calf serum (FCS), 100 units/ml penicillin, 100 µg/ml streptomycin, 2 mM L-glutamine, 1 mM sodium pyruvate, and 1x non-essential amino acids. HEK293T (human, female, kidney; ACC-635, DSMZ, RRID: CVCL_0063) cells were grown in DMEM with supplementation of 10% FCS, 100 units/ml penicillin, 100 µg/ml streptomycin, 2 mM L-glutamine. All cells were grown at 37°C in a 5% CO2 humidified incubator.

### Preparation of pseudotyped particles

Expression plasmids for vesicular stomatitis virus (VSV, serotype Indiana) glycoprotein (VSV-G) and SARS-2-S variants B.1.1.7, B.1.351 and B.1.617 (codon-optimized; with a C-terminal truncation of 18 amino acid residues) have been described elsewhere (24, 42). Transfection of cells was carried out by Transit LT-1 (Mirus). Rhabdoviral pseudotype particles were prepared as previously described (43). A replication-deficient VSV vector in which the genetic information for VSV-G was replaced by genes encoding two reporter proteins, enhanced green fluorescent protein and firefly luciferase (FLuc), VSV∗ΔG-FLuc (44) (kindly provided by Gert Zimmer, Institute of Virology and Immunology, Mittelhäusern, Switzerland) (Berger Rentsch and Zimmer, 2011) was used for pseudotyping. One day after transfection of HEK293T cells to express the viral glycoprotein, they were inoculated with VSV∗ΔG-FLuc and incubated for 1-2 h at 37°C. Then the inoculum was removed, cells were washed with PBS and fresh medium added. After 16-18 h, the supernatant was collected and centrifuged (2,000 × g, 10 min, room temperature) to clear cellular debris. Cell culture medium containing anti-VSV-G antibody (I1-hybridoma cells; ATCC no. CRL-2700) was then added to block residual VSV-G-containing particles. Samples were then aliquoted and stored at -80°C.

### Pseudovirus neutralisation assay

For pseudovirus neutralisation experiments, VeroE6 were seeded in 96-well plates one day prior (6,000 cells/well). Heat-inactivated (56°C, 30 min) sera were serially titrated in PBS, pseudovirus stocks added (1:1, v/v) and the mixtures incubated for 30 min at 37°C before being added to cells. After an incubation period of 16-18 h, transduction efficiency was analyzed. For this, the supernatant was removed, and cells were lysed by incubation with Cell Culture Lysis Reagent (Promega) at room temperature. Lysates were then transferred into white 96-well plates and FLuc activity was measured using a commercially available substrate (Luciferase Assay System, Promega) and a plate luminometer (Orion II Microplate Luminometer, Berthold). For analysis of raw values (RLU/s), background signal of an uninfected plate was subtracted and values normalized to pseudovirus treated with PBS only. Results are given as serum dilution resulting in 50% virus neutralization (NT50) on cells, calculated by nonlinear regression ([Inhibitor] vs. normalized response -- Variable slope) in GraphPad Prism Version 9.1.1.

### Determination of CD4^+^ and CD8^+^ T SARS-CoV-2 spike -specific cell responses by intracellular cytokine staining (ICS)

Cryopreserved PBMCs of study participants were thawed and rested overnight at 37°C with 1 µl/ml of DNAse (DNase I recombinant, RNase-free (10,000 U) Roche), in RPMI medium supplemented to contain a final concentration of 10% FCS (Corning Life Sciences/Media Tech Inc, Manassas, VA), 10 mM HEPES, 1x MEM nonessential amino acids (Corning Life Sciences/Media Tech Inc, Manassas, VA), 1 mM Sodium Pyruvate (Lonza, Walkersville, MD, U.S.A), 1mM Penicillin/Streptomycin (Pan Biotech, Germany) and 1x 2-Mercaptoethanol (GIBCO, Invitrogen, Carlsbad, CA, U.S.A). Stimulation of PBMCs for detection of cytokine production by T cells was adapted from Kasturi *et al*., 2020 (45). Briefly, 1×10^6^ PBMCs were cultured in 200 μl final volume in 96-well U bottom plate in the presence of anti-CD28 (1 μg/ml) and anti-CD49d (1 μg/ml) [Biolegend] under the following conditions: a) negative control with DMSO, b) SARS-CoV-2 spike peptide pools (1-315 peptides from Wuhan-Hu-1, B.1.1.7 (Alpha), B.1.351 (Beta) and P.1 (Gamma) SARS-CoV-2 spike, JPT Germany) at a final concentration of 2 μg/ml, c) PMA/Ionomycin, d) CEFX Ultra Super Stim peptide pool (176 peptide epitopes for a broad range of HLA subtypes and different infectious agents, JPT Germany) at a final concentration of 2 μg/ml. Cells were cultured for 2 hours before adding Brefeldin A at 10 μg/ml (Sigma-Aldrich, St Louis, MO) for an additional 5 hours. Cells were then washed with PBS, and stained for dead cells (Live/ Dead Fixable; Aqua from Thermo Fisher) in PBS at room temperature for 10 minutes. Without washing, cells were incubated with surface antibody cocktail (prepared in 1:1 of FACS buffer and brilliant staining buffer) for 30 minutes at room temperature with BV510- anti-human CD14 (clone M5E2), BV510-anti-human CD19 (clone HIB19), AF700 anti-human CD3 (clone OKT3), BV605 CD4 (clone OKT4), PerCP-Cy5.5 CD8 (clone RPA-T8) from Biolegend. Next, cells were fixed using Cytofix/Cytoperm buffer (BD Biosciences, CA) for 20 minutes at room temperature, and then kept in FACS buffer at 4°C overnight. 1x Perm/Wash (BD Biosciences, CA) was used for cells permeabilization for 10 minutes at room temperature and followed by intracellular staining for 30 minutes at room temperature with AF647 anti-human IFNγ (clone 4S.B3) and AF488 anti- human IL-2 (clone MQ1-17H12) from Biolegend, and PE/Cy7 anti- human TNFα (clone Mab11) from Thermo Fisher Scientific. Up to 100,000 live CD3^+^ T cells were acquired on a LSRFortessa flow cytometer (BD Biosciences), equipped with FACS Diva software. Analysis of the acquired data was performed using FlowJo software (version 10.7.1). The background from each participant was removed by subtracting the % of spike^+^ cells to the % of DMSO^+^ cells. An arbitrary value below 0·01% of CD4^+^/CD8^+^ T cells was considered negative.

### Statistical analysis

The SARS-CoV-2 convalescent individual was excluded in all statistical analyses. Non-parametric Spearman rank correlation was used to check for possible associations at single blood sample measurements. A paired t-test was used to compare the adverse event scores calculated for each participant after both vaccinations. For this, the individual mean differences were checked for normal distribution by means of QQ-plots and histograms. A comparison of participants receiving heterologous vaccination with controls who received homologous BNT162b2 vaccinations after the last blood sample measurements was done by the Mann-Whitney-U test because of skewed distributions for neutralization scores as well as IgM/IgG measurements. Longitudinal antibody measurements were analyzed by means of a mixed linear regression model including a random intercept in order to account for the repeated measures structure of the underlying data. The mixed linear model approach enabled to simultaneously account for possible confounding due to participants’ sex and for the presence of missing data (46). Therefore, no formal imputation of missing interim values was required. A two-sided alpha error of 5% was applied to analyses. Analysis of repetitive measurements of sera provided by a cohort of 26 participants can be considered statistically valid. T cell reactivity before vaccination and 72-73 days after prime (2 weeks after boost) or between variants was compared with Wilcoxon matched-pairs signed rank test. All analyses were done by GraphPad Prism version 9.1.1 for Windows, GraphPad Software, San Diego, California USA, www.graphpad.com, R (version 4.0.1) and SAS (version 9.4).

## Data Availability

Data is available upon request after positive peer-review evaluation.

## Acknowledgments

We thank all participants for regular blood donations. We thank Nicola Schrott, Regina Burger, Jana Romana Fischer, Birgit Ott, Carolin Ludwig, Katrin Ring, Nadine Pfeifer, Maxine Rustler, Vivien Prex for skillful laboratory assistance. We thank Sarah Warth, Simona Ursu, and Christian Buske of the flow cytometry core facility for support with flow cytometric analysis. We thank the Robert Koch Institute (RKI) for financial support. This project has received funding from the European Union’s Horizon 2020 research and innovation programme under grant agreement No 101003555 (Fight-nCoV) to J.M., the German Research Foundation (CRC1279) to F.K. and J.M., the BMBF to F.K. (Restrict SARS-CoV-2) and an individual research grant (to J.A.M.). J.A.M. is indebted to the Baden-Württemberg Stiftung for the financial support of this research project by the Eliteprogramme for Postdocs. R.G., A.S., and C.C. are part of the International Graduate School in Molecular Medicine Ulm. S.P. received funding from BMBF (01KI1723D, 01KI20328A, 01KI20396, 01KX2021) and the county of Lower Saxony (14-76103-184, MWK HZI COVID-19). H.S. acknowledges funding from the Ministry for Science, Research and the Arts of Baden-Württemberg, Germany and the European Commission (HORIZON2020 Project SUPPORT-E, no. 101015756).

## Author contributions

Conceptualization, J.A.M., J.M., R.G., M.Z., A.S.; Funding acquisition, J.A.M., J.M., F.K., S.P., H.S.; Investigation, R.G., M. Z., A.S., C.C., A.G., D.K., S.E.E., A.B. J.K.; Essential resources, B.M., M.H., S.P., B.J., H.S., F.K., J.M., J.A.M.; Writing, J.A.M., R.G., M.Z., A.S.; Review and editing, all authors. R.G., M.Z., A.S. contributed equally. R.G., M.Z., A.S. and J.A.M verified the underlying data.

## Data and materials availability

Raw data is available upon request.

