## Supplementary Appendix for "Heterologous ChAdOx1 nCoV-19 and BNT162b2 prime-boost vaccination elicits potent neutralizing antibody responses and T cell reactivity"

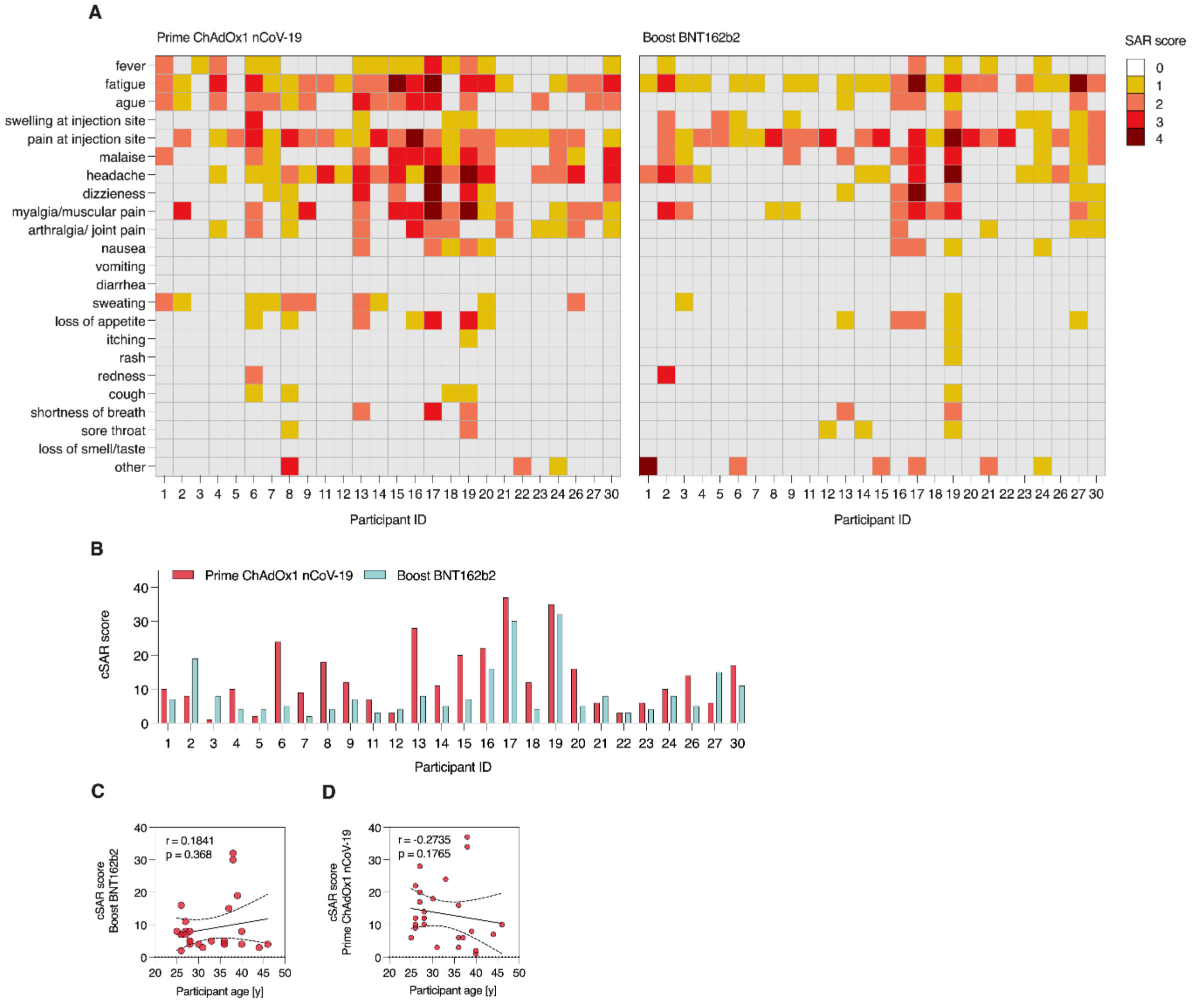


**Figure S1. Extended analysis of solicited adverse reactions following ChAdOx1 nCoV-19 prime and BNT162b2 boost vaccination.** (A) Heatmap showing SAR scores per participant and symptom used for calculation of cSAR scores. Severity is graded on a scale of 1-2 (for some symptoms) or 1-3 (for most), according to Common Terminology Criteria for Adverse Events (US Department of Health and Human Services, Version 4). For calculation of cSAR score (A, B), symptom gradings are summed and an additional score point is added for symptoms lasting more than 24 h (final scores 1-4 per symptom). Correlation analysis for cSAR scores with participant age for prime (C) and boost (D) vaccination.

**Appendix Table 1. Serum donors vaccinated twice with BNT162b2**

| | **Total** | **m** | **f** |
|---|---|---|---|
| Participants | 15 | 11 | 4 |
| Age median | 41 (25-65) | 41 (32-65) | 38 (25-55) |
| Prior SARS-CoV-2 infection | 0 | 0 | 0 |

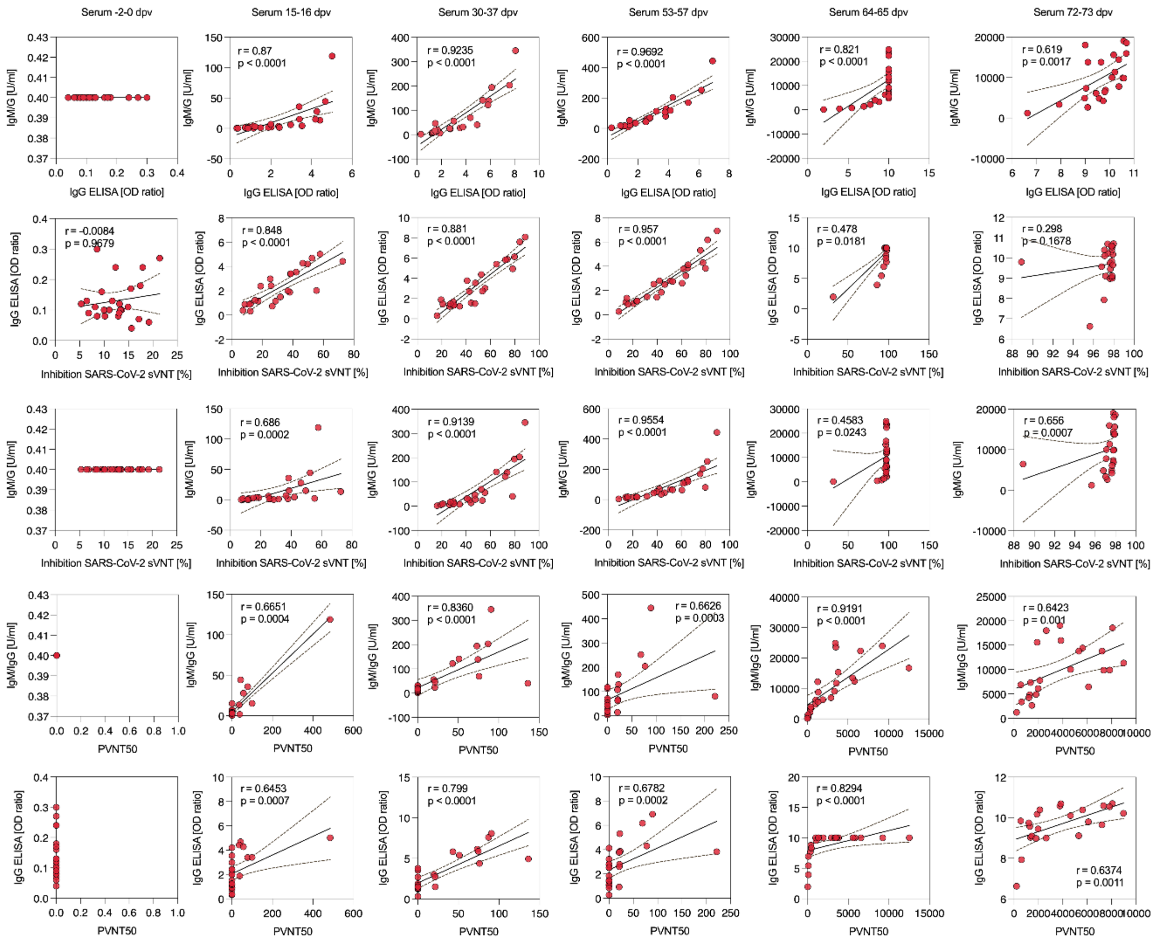


**Figure S2. Correlation analysis of humoral response metrics.** Data on humoral response (IgG, IgA, IgM/G, PVNT50, Inhibition SARS-CoV-2-sVNT) were analysed for correlation for each timepoint. Spearman correlation, two-tailed p values, dashed lines indicate 95% confidence interval. The SARS-CoV-2 convalescent individual was excluded from the analysis.

**Appendix Table 2. Summary of correlation analysis of humoral response metrics.** Spearman r values of Figure S2.

| | **-2 - 0** | **15 – 16** | **30 - 37** | **53 - 57** | **64 - 65** | **72 - 73** |
|---|---|---|---|---|---|---|
| **IgG:IgM/G** | / | 0·87 | 0·9235 | 0·9692 | 0·821 | 0·619 |
| **sVNT:IgG** | -0·0084 | 0·848 | 0·881 | 0·957 | 0·478 | 0·296 |
| **sVNT:IgM/G** | / | 0·686 | 0·9139 | 0·9554 | 0·4583 | 0·656 |
| **PVNT50:IgM/G** | / | 0·6651 | 0·836 | 0·6626 | 0·9191 | 0·6423 |
| **PVNT50:IgG** | / | 0·6453 | 0·799 | 0·6782 | 0·8294 | 0·6374 |

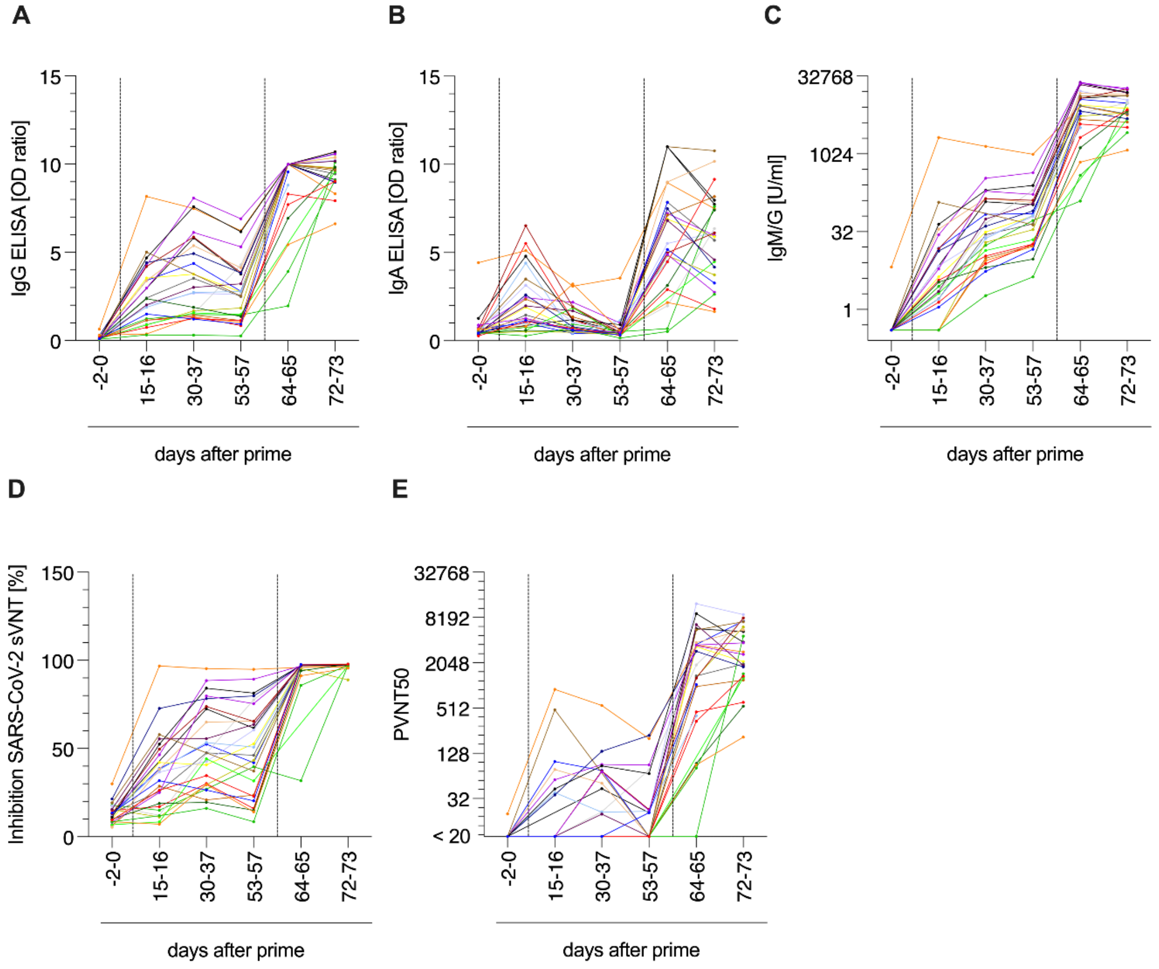


**Figure S3. Time course of humoral responses.** Time course of anti-SARS-CoV-2 S1 spike domain (A) IgG and (B) IgA titres. (C) Time course of anti-SARS-CoV-2 spike IgG and IgM responses as units per ml (U/ml) by immunoassay. (D) Time course of SARS-CoV-2 surrogate virus ACE2-RBD interaction neutralization as assessed by surrogate virus neutralisation test (sVNT). (E) VSV-based B.1.1.7 SARS-CoV-2 spike pseudovirus neutralization assay. Titres expressed as serum dilution resulting in 50% pseudovirus neutralization (PVNT50).

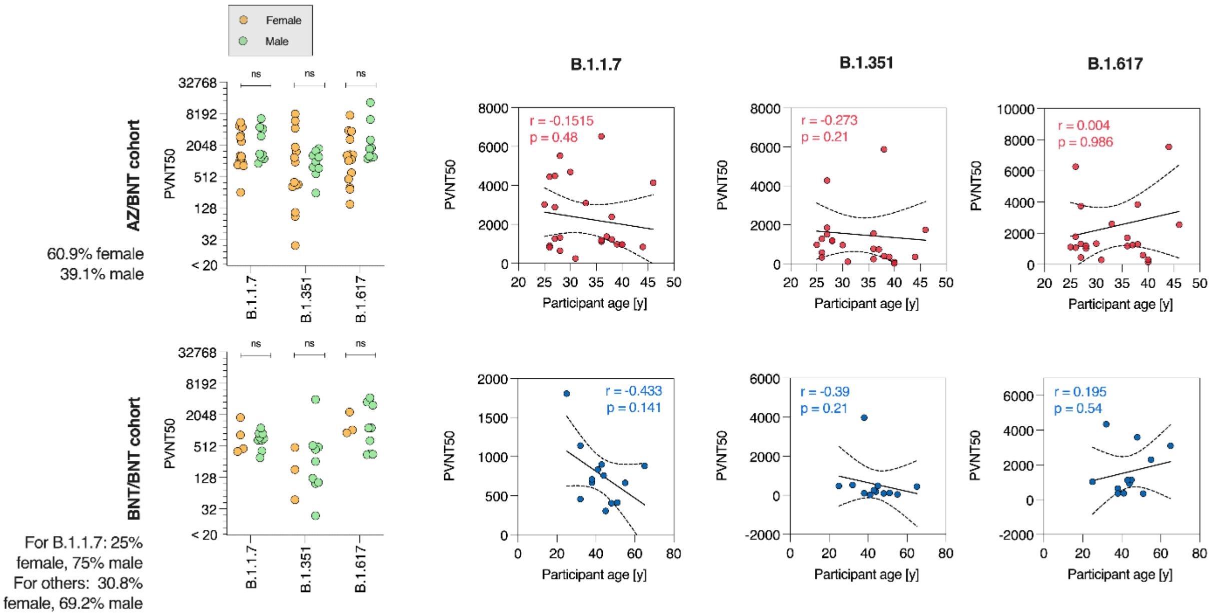


**Figure S4. Analysis of sex and age correlation with neutralizing antibody titres (PVNT50) in heterologous and homologous cohort.** Neutralizing antibody titres (PVNT50) were clustered according to participant sex (left) and analysed for correlation with participant age (right) for each of the variants tested within the study. Spearman correlation, two-tailed p values, dashed lines indicate 95% confidence interval. Wilcoxon matched-pairs signed rank test compares PVNT50 between male/female participants. ns = not significant

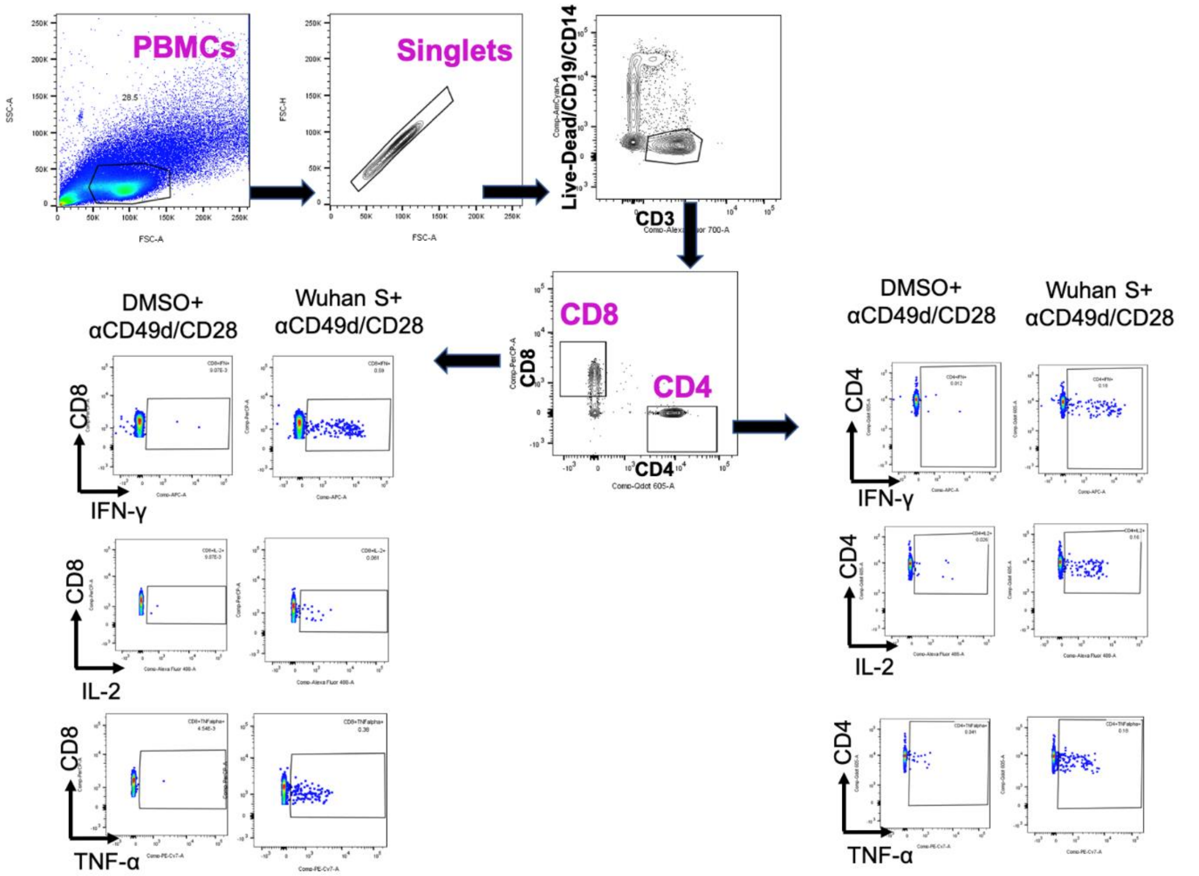


**Figure S5. Gating strategy for analysis of T cell reactivity.** SARS-CoV-2 spike peptide stimulated and unstimulated (DMSO) PBMCs were initially gated on the basis of light scatter (SSC-A versus FSC-A) and for singlets (FSC-H versus FSC-A). Dead cells, monocytes, and B cells were excluded using a dump channel and by gating on $CD3^+$ T cells. Total $CD8^+$ and $CD4^+$ cells were then selected, and individual cytokine gating was performed.

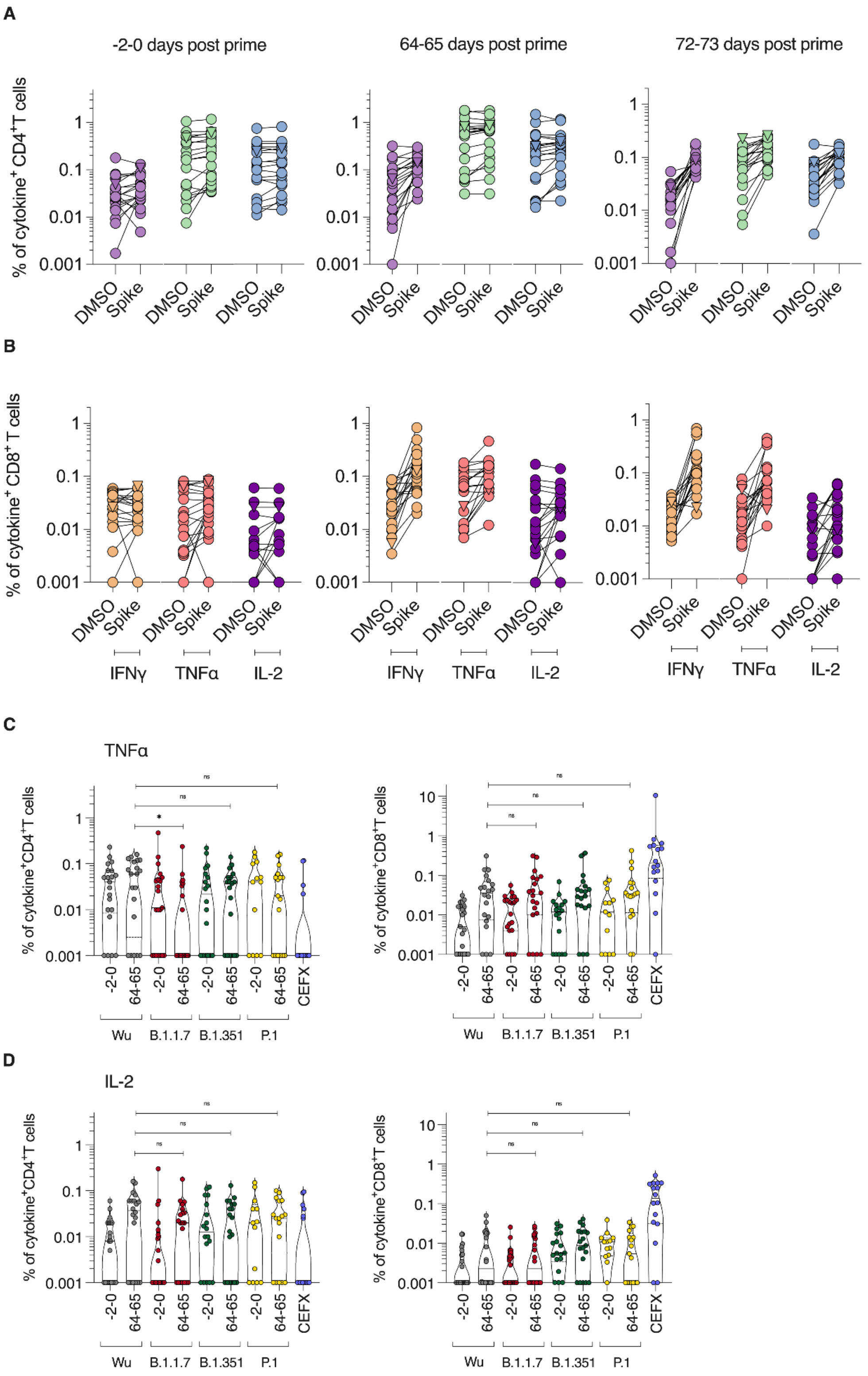


**Figure S6. CD4$^{+}$ and CD8$^{+}$ T cell responses to SARS-CoV-2 spike peptide pools or DMSO (mock).** PBMCs from day -0-2 pre- and day 64-65 and 72-73 post prime vaccination (1-2 weeks post boost) of study participants were stimulated with SARS-CoV-2 spike peptide-pools or DMSO (mock) and cytokine production determined by flow cytometry. Reactivity of (A) CD4$^{+}$ or (B) CD8$^{+}$ T cells to SARS-CoV-2 Wuhan-Hu-1 spike peptides. Production of (C) TNFα or (D) IL-2 upon stimulation with Wuhan-Hu-1 (Wu; data from Fig. 3A), B.1.1.7, B.1.351, P.1 peptide pools or peptide pools of epitopes of different infectious agents (CEFX). Triangle symbol indicates SARS-CoV-2 convalescent individual, where cytokine release was already high in absence of stimulation. Wilcoxon matched-pairs signed rank test compares cytokine-positive cells at 64-65 weeks after prime (1 week after boost) upon stimulation with different SARS-CoV-2 spike variants, *** $p < 0·0001$, * $p < 0·05$.
